# Emerging Links Between COVID-19 and Cardiovascular & Cerebrovascular Thromboembolic Events: A Systematic Review

**DOI:** 10.1101/2023.09.05.23295067

**Authors:** Abhimanyu Agarwal, Binay K Panjiyar, Patel Dhwani Manishbhai, Monica Ghotra, Mahato Gulam Husain Nabi Husain, Upadhyay Ronak Brijeshkumar

## Abstract

COVID-19, caused by the SARS-CoV-2 virus, initially identified as a respiratory illness, has increasingly been linked to a broader range of organ complications. This systematic review explores the impact of COVID-19 on cardiovascular and cerebrovascular health, focusing on thromboembolic events in post-COVID patients. A comprehensive literature search was conducted in PubMed and Google Scholar databases up to July 2023, utilizing the Preferred Reporting Items for Systematic Reviews and Meta-Analyses (PRISMA) guidelines. Studies meeting eligibility criteria were analyzed for outcomes and associations between COVID-19 and cardiovascular and cerebrovascular events. The review includes 6 studies involving over 12 million patients, demonstrating a strong connection between COVID-19 and elevated risks of cardiovascular and cerebrovascular thromboembolic events. The risk of these events is evident in conditions such as ischemic heart disease, stroke, and cardiac arrhythmias. The burden of these events beyond the acute phase of the disease is concerning, warranting further exploration of long-term implications. Variability in event rates among different cohorts and healthcare settings underscores the need for understanding underlying factors influencing these differences. Potential mechanisms behind these events include endothelial dysfunction, systemic inflammation, and viral invasion. Implications for public health policies, clinical guidelines, and future research directions are discussed. This review serves as a valuable resource for healthcare providers, policymakers, and researchers to enhance patient care, outcomes, and preparedness for future waves of COVID-19 infections. However, there remain unexplored aspects of the COVID-19 and thromboembolic events relationship, urging further investigations into mechanistic insights and potential therapeutic interventions.

## INTRODUCTION

COVID-19 is primarily known as a respiratory illness caused by the Severe Acute Respiratory Syndrome - CoronaVirus -2 (SARS-CoV-2). However, growing clinical reports and research studies have highlighted the virus’s propensity to affect various organs beyond the respiratory system.

As the initial wave of infections has receded, an increasing body of evidence points towards the significant impact of COVID-19 on cardiovascular and cerebrovascular health. An emerging concern is the potential development of acute and chronic cardiovascular and cerebrovascular events in patients who have recovered from the acute phase of the disease.

The potential mechanisms behind the development of cardiovascular and cerebrovascular events in post-COVID patients remain elusive but are believed to be multifactorial. Systemic inflammation, endothelial dysfunction, microvascular thrombosis, and direct viral invasion are among the suspected mechanisms[1-9] contributing to these complications. Understanding these pathways is crucial for devising appropriate preventive strategies and targeted treatment modalities.

This systematic review will also address the implications of post-COVID cardiovascular and cerebrovascular events on the overall burden of disease, healthcare resources, and patient management. Moreover, it will highlight gaps in current knowledge and potential areas of future research to guide the medical community in further understanding and addressing this emerging health challenge. As the world continues to grapple with the aftermath of the COVID-19 pandemic, the findings of this systematic review can serve as a valuable resource for healthcare providers, policymakers, and researchers to optimize patient care, improve outcomes, and enhance preparedness for potential future waves of infections.

## MATERIALS AND METHODS

PubMed and Google Scholar databases were systematically searched from inception until 31 July 2023 and a systematic review of the thromboembolic cardiovascular and cerebrovascular events in post COVID19 patients was conducted by following methods outlined in the Preferred Reporting Items for Systematic Reviews and Meta-Analyses (PRISMA)[10]. Institutional review board approval was not sought because of the use of publicly available data.

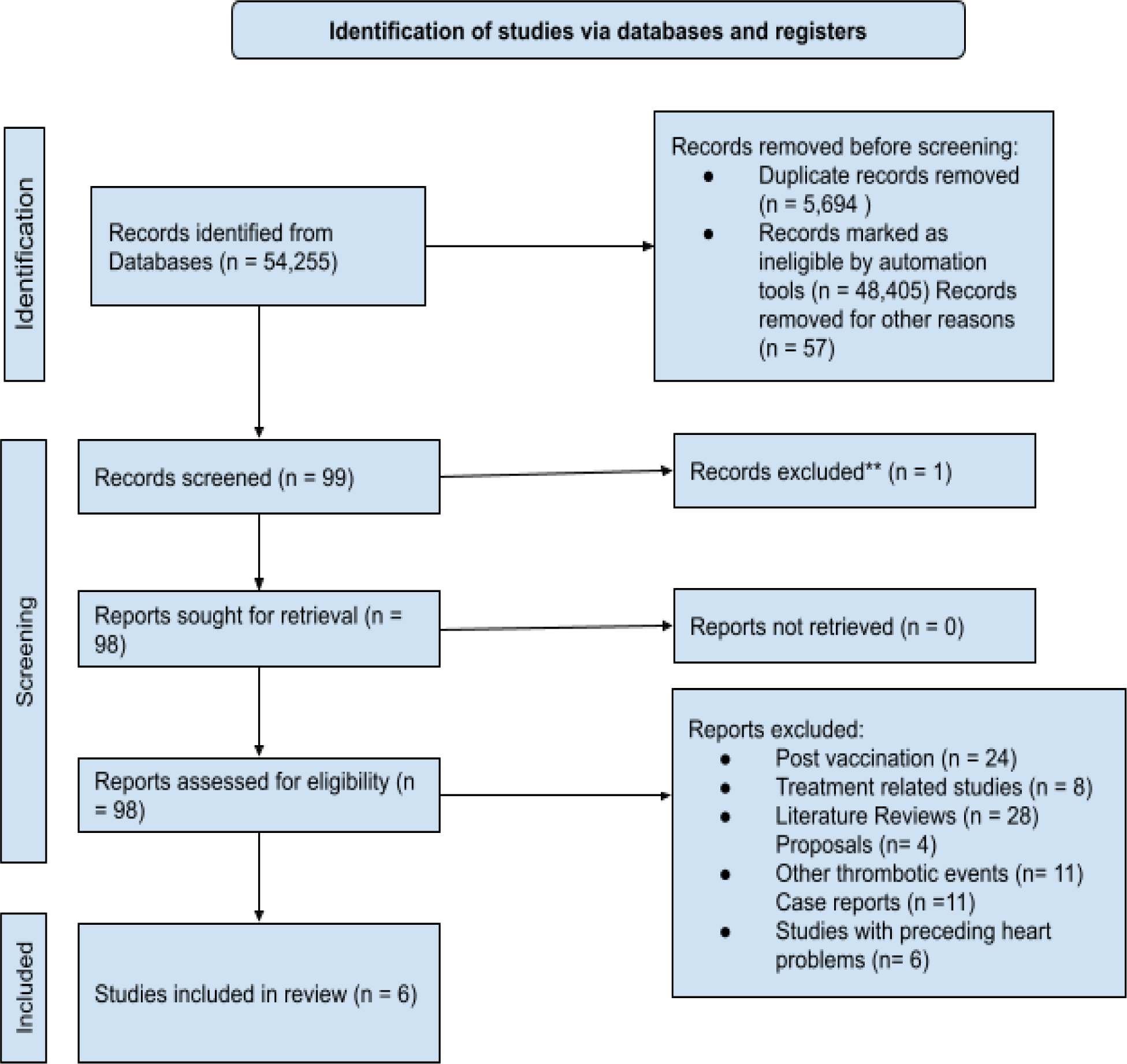

### Eligibility Criteria

Studies published in English-language only were considered eligible if they meet following criteria:

| Inclusion criteria: | Exclusion criteria: |
| --- | --- |
| 1. All Free papers<br>2. Observational studies, randomized controlled trials.<br>3. All participants with past medical history of COVID19 infection.<br>4. Only human subjects were included.<br>5. Outcome of interest, i.e. COVID19 patients with cardiovascular or cerebrovascular thromboembolic events, was clearly reported via recognized scoring system | 1. All paid papers, Animal studies and non-english text<br>2. Studies with patients status post vaccination.<br>3. Studies discussing pathophysiology of thromboembolic events<br>4. Did not report data about thromboembolic events or only reported biomarkers or genetic markers.<br>5. Did not present original data, or were editorials, case reports, case series, systematic reviews or meta analysis. |

### Search Strategy

The population, intervention/condition, control/comparison, and outcome (PICO) criteria were utilized to conduct a thorough literature review. The search was conducted on databases such as PUBMED (including Medline) and Google Scholar Libraries, using relevant keywords, ‘cardiovascular’, ‘cerebrovascular’, ‘thromboembolic’, ‘embolic’, ‘thrombotic’, ‘post COVID19’, ‘SARS CoV 2’ and ‘blood clotting’. The medical subject heading (MeSH) approach for PubMed (including Medline) and Google Scholar, as detailed in Table 2, was employed to develop a comprehensive search strategy.

**TABLE 2.** Showing the Search strategy, search Engines used, and the number of results displayed.

| Database | Search strategy | Search Results |
| --- | --- | --- |
| Pubmed | ((((((((((((((((((Cardiovascular[Title/Abstract]) OR (cardio-vascular[MeSH Terms])) OR (abnormalities, heart[MeSH Terms])) OR (vascular[MeSH Terms])) AND (cerebrovascular[MeSH Terms])) AND (thrombotic[MeSH Terms])) OR (embolic[MeSH Terms])) OR (thromboembolic[MeSH Terms])) OR (blood clotting[MeSH Terms]) AND (events[MeSH Terms])) OR (abnormalities[MeSH Terms])) OR (episodes[MeSH Terms])) OR (incidents[MeSH Terms])) AND (post COVID19[MeSH Terms])) OR (coronavirus[MeSH Terms])) OR (coronavirus, sars[MeSH Terms])) OR (coronavirus, sars associated[MeSH Terms])) OR (coronavirus, sars related[MeSH Terms])) OR (SARSCov2[MeSH Terms])) OR (long term covid[MeSH Terms])) OR (long covid[MeSH Terms])) ) AND (("2013/01/01"[Date - Publication] : "2023/08/01"[Date - Publication])) | 29,955 |
| Google Scholar | cardiovascular cerebrovascular events in post covid patients<br>Thrombotic OR Thromboembolic OR Embolic | 24300 |

### Study Identification

Investigators screened articles by title and abstract using predefined inclusion and exclusion criteria and a standardized data form. If the article did not meet inclusion criteria based on the abstract, the full text was not reviewed. The inclusion of full text was decided by consensus. All results were exported to EndNote, an open-source research tool, for organizing and analyzing data, where duplicates were eliminated.

### Data Extraction and Outcomes

A structured data collection form was employed to gather information from each study, including study design, patient characteristics, baseline variables, thromboembolic event ascertainment, maximum adjusted covariates and hazard risks (HRs) with 95% CIs. The study encompassed all diagnoses of thromboembolic events, including cardiovascular and cerebrovascular cases. We focused our review on the association between COVID19 and thromboembolic events.

### Risk of Bias Assessment

The Newcastle–Ottawa Scale (NOS) items and The Cochrane Collaboration’s tool for assessing risk of bias in randomized trials were employed to evaluate the quality of the article. NOS score ≥ 6 stars were defined as high quality, and NOS score < 6 stars as low quality

## RESULTS

### Impact of COVID19 on cardiovascular thromboembolic events

Three studies[11,12,13] involving 12,419,154 patients discussed cardiovascular thromboembolic events among post COVID19 patients.

In a study by Xie et al.[11], various cardiovascular conditions were assessed for their risks (HR) and burdens (per 1000 persons at 12 months). The risks and burdens of a composite of these dysrhythmia outcomes were (1.69; 95% CI: 1.64, 1.75) and (19.86; 95% CI: 18.31, 21.46) respectively, for inflammatory diseases of the heart or pericardium were (2.02; 95% CI: 1.77, 2.30) and (1.23; 95% CI: 0.93, 1.57), for ischemic heart disease outcomes being (1.66; 95% CI: 1.52, 1.80) and (7.28; 95% CI: 5.80, 8.88) and for other cardiovascular disorders were (1.72; 95% CI: 1.65, 1.79) and (12.72; 95% CI: 11.54, 13.96). MACE (HR: 1.55; 95% CI: 1.50, 1.60) included MI, stroke, and all-cause mortality, while any cardiovascular outcome (HR: 1.63; 95% CI: 1.59, 1.68) encompassed pre-specified cardiovascular events from the study.

According to Ortega-Paz et al.[12], after adjusting for various factors, Patients within the COVID-19 cohort exhibited a notably increased incidence of all-cause death (HR: 2.82; 95% CI: 1.99, 4.00; p=0.001) and serious cardiac arrhythmias (HR: 3.37; 95% CI: 1.35, 8.46; p=0.010) in comparison to the control cohort, with no significant variance in MI rates (HR: 1.44; 95% CI: 0.64, 3.26; p=0.383). In the realm of exploratory endpoints, the COVID-19 cohort demonstrated a higher frequency of adverse cardiovascular events (HR: 2.63; 95% CI: 1.75, 3.96; p=0.001) in contrast to the control cohort, and although a numerical increase in MACE was observed in the COVID-19 cohort compared to the control (HR: 1.74; 95% CI: 0.97, 3.13; p=0.065), this did not reach statistical significance.

After accounting for cohort variations using the propensity score, Ward et al.[13] observed no significant distinction in the risk of myocardial infarction (HR: 0.93; 95% CI: 0.85, 1.03) between the COVID-19 and influenza cohorts.

### Impact of COVID19 on cerebrovascular thromboembolic events

Six studies[11-16] involving 12,452,321 patients discussed cerebrovascular thromboembolic events among post COVID19 patients.

Xie et al.[11] found that Individuals who managed to survive the initial 30 days following COVID-19 infection displayed an elevated likelihood of experiencing a stroke (HR:1.52; 95% CI: 1.43, 1.62) and a burden of 4.03 per 1,000 individuals at the 12-month mark. Additionally, there was an increased risk of TIA (HR: 1.49; 95% CI: 1.37, 1.62). The combined risk associated with these various cerebrovascular outcomes were (1.53; 95% CI: 1.45, 1.61).

Regarding the secondary endpoints, after adjusting for various factors, Ortega-Paz et al.[12] observed that individuals within the COVID-19 group exhibited a greater occurrence of all-cause mortality (HR: 2.82; 95% CI: 1.99, 4.00; p=0.001). No notable distinctions between the cohorts were evident in terms of stroke rates (HR: 3.27; 95% CI: 0.77, 13.90; p=0.109). When focusing on exploratory endpoints, the COVID-19 cohort displayed a higher incidence of ischemic stroke (0.7% vs. 0.0%; p=0.017).

After accounting for cohort variations using the propensity score, Ward et al.[13] observed no significant distinction in the risk of stroke (HR: 1.11; 95% CI: 0.98, 1.25) or myocardial infarction (HR: 0.93; 95% CI: 0.85, 1.03) between the COVID-19 and influenza cohorts.

Siegler et al.[14] reported that Out of a total of 14,483 individuals who tested positive for SARS-CoV-2 in laboratory tests, 172 were found to have an acute cerebrovascular event (CVE) incidence rate of CVE was 1,130 per 100,000 patients (95% CI: 970, 1320 per 100,000). The rates of CVE varied across different medical sites, ranging from 0.19% to 5.04%, and there was an observable trend where sites treating higher numbers of COVID-19 patients experienced lower rates of CVE and acute ischemic stroke. The most frequent type of CVE observed was acute ischemic stroke, affecting 156 patients (1.08% of the cohort), 28 patients have primary intracerebral hemorrhage (0.19% incidence rate) and 3 had cerebral venous sinus thrombosis (0.02% incidence rate). Fourteen patients (8.1%) experienced both acute stroke and intracerebral hemorrhage.

According to Sabayan et al.[15], a total of 15 out of 18,407 COVID-19 cases (0.81 per 1000 cases, 95% CI: 0.49-1.3) were identified as having cerebrovascular events. The time interval between the onset of systemic manifestations and the development of neurological symptoms varied from one to 16 days, with a median interval of 7 days. Among the cerebrovascular events, 14 patients experienced acute ischemic stroke, while one patient had a subarachnoid hemorrhage. A majority of patients (73%) had underlying cardiovascular risk factors, and all patients underwent TTE, with no indications of myocarditis, endocarditis, or pericarditis. Cardiac monitoring did not reveal atrial fibrillation or other arrhythmias. Elevated levels of CRP, indicative of a pro-inflammatory state, were observed in all tested patients. Doppler ultrasound of cervical arteries showed no significant stenosis in four ischemic stroke patients. Stroke severity varied, with 13% having mild severity (NIHSS ≤6), 40% moderate severity (NIHSS: 7-12), and 47% severe severity (NIHSS ≥13). One patient received intravenous tissue plasminogen activator (IV-tPA) with a positive response. Unfortunately, the mortality rate was 40%, and among the survivors, significant disability (mRS>2) was present in all but one patient.

Among individuals with COVID-19, Dhamoon et al.[16] found that the initial severity of strokes was notably higher compared to those without COVID-19, as indicated by a mean NIH Stroke Scale of 15.5 vs. 9.6. Although the distribution of stroke subtypes was comparable between the two cohorts (p=0.45), the underlying causes of ischemic stroke differed significantly. Notably, 51.8% (n=43) of COVID-19-positive patients had a cryptogenic etiologic subtype, contrasting with only 22.3% (n=27) among the COVID-19-negative cohort (p<0.0001). Additionally, COVID-19 patients experienced lower incidences of small-vessel ischemic strokes (6.0%, n=5) and higher occurrences of cardioembolic ischemic strokes (28.9%, n=24), in contrast to non-COVID-19 patients with rates of 17.4% (n=21) and 42.2% (n=51) respectively. Notably, COVID-19-positive patients exhibited a more frequent incidence of ischemic strokes in regions such as the temporal (p=0.04), parietal (p=0.002), occipital (p=0.002), and cerebellar (p=0.027) areas, in comparison to COVID-19-negative patients. However, there were no discernible differences in the location of intracerebral hemorrhages between the two groups.

## DISCUSSION

During our review of 6 studies involving 12,452,321 patients, we found a strong association of COVID19 with cardiovascular and cerebrovascular thromboembolic events.

Among the analyzed patients by Szegedi et al.[17], there were 19 instances of hemorrhagic stroke (HS), with four of those cases presenting SAH, and an additional patient experiencing HS followed by acute ischemic stroke (AIS). TIA was observed in six patients, while the majority suffered from AIS, numbering 170 cases. The initial NIHSS score upon admission indicated severity, with a median score of 16 (IQR: 10–22). Out of 198 patients with available medical histories, merely 29 exhibited no prior chronic illnesses as risk factors; the rest had prevalent conditions such as hypertension, diabetes mellitus, and hyperlipidemia. Concerning outcomes, data was limited and attainable for only 116 patients: 74 patients succumbed (64%), 23 had unfavorable outcomes (19%), and merely 19 demonstrated favorable recoveries (16%). Detailed functional outcomes were absent in some instances, wherein 11 patients were moved to rehabilitation and seven were discharged home without neurological status descriptions. In cases involving neurological interventions (thrombolysis and/or mechanical thrombectomy), documented in a subset of only 30 cases, the proportion of favorable outcomes closely resembled those with conservative treatment (17%); mortality rate was 48%, and 35% experienced unfavorable outcomes. In 7 out of 30 intervention cases, outcomes were ambiguously defined as transfer to rehabilitation or home.

In a collective analysis of 16 studies encompassing 4448 patients by Pranata et al.[18], the presence of cerebrovascular disease exhibited a significant association with an elevated risk of adverse composite outcomes (RR: 2.04; 95% CI: 1.43, 2.91; p<0.001). This relationship was further substantiated by subgroup analysis, which demonstrated an increased risk of mortality (RR: 2.38; 95% CI: 1.92, 2.96; p<0.001) linked to cerebrovascular disease. While the association between cerebrovascular disease and severe COVID-19 was borderline significant (RR: 1.88; 95% CI: 1.00, 3.51; p=0.05), cardiovascular disease similarly displayed a strong correlation with adverse composite outcomes (RR: 2.23; 95% CI: 1.71, 2.91; p<0.001). Subgroup analysis unveiled heightened risks of mortality (RR: 2.25; 95% CI: 1.53, 3.29; p<0.001) and severe COVID-19 (RR: 2.25; 95% CI: 1.51, 3.36; p<0.001) in individuals with cardiovascular disease. Notably, meta-regression analyses for both cerebrovascular and cardiovascular diseases demonstrated that these associations remained consistent across various factors such as gender, age, hypertension, diabetes, cardiovascular diseases, and respiratory comorbidities (all p-values >0.05).

The multifaceted nature of the probable processes behind the development of cardiovascular and cerebrovascular events in post-COVID patients is yet unknown. Dwiputra et al.[19] conducted a study aimed at uncovering the diverse pathogenesis mechanisms. Endothelial dysfunction, identified as a crucial element of COVID-19, holds significant prognostic importance due to its correlation with clinical complications and unfavorable results. Regarding IL-6, it has previously been established as a potent indicator of increased COVID-19 severity. Additionally, there have been extensive reports on the regulatory role of IL-6 in the initiation and progression of atherosclerosis, suggesting a potential shared pathogenetic pathway between COVID-19 and cardiovascular diseases replicated in study conducted by Oikonomou et al.[20], Ridker et al.[21], and Cruz et al.[22]. Specifically, patients admitted to the ICU and non-survivors exhibited notably higher IL-6 levels as compared to others.

The presence of diffuse endothelial damage in the alveolar epithelium and vascular microthrombi, along with elevated PaCO2 levels, were observed in COVID-19 patients, as reported by Spadaro et al.[23]. In Ambrosino et al.’s research[24] patients who had COVID-19 and were referred for pulmonary rehabilitation, endothelial function remained impaired even after two months of hospitalization. The study by Meshref et al.[25] and Nauen et al.[26] suggested the involvement of thrombocytosis and megakaryocyte-related thrombosis based on increased platelet levels.

Kihira et al.[27] discovered that SARS-CoV-2 primarily targets the major blood vessels in the circle of Willis. with 31.7% of COVID-19 patients experiencing large vessel occlusion, while 5.9% had small vessel occlusion. Potential mechanisms include vascular restructuring, direct or immune-triggered harm to endothelial cells, disruption of natural fibrinolysis processes, and heightened blood coagulation tendencies[28,29]. Studies have revealed that the majority of individuals from whom SARS-CoV-2 proteins/mRNA were derived from their cerebrospinal fluid exhibited indications indicative of encephalitis or demyelinating conditions, with respiratory symptoms being a rarity. Additionally, these proteins were detected in the brain, olfactory bulb/nerve or olfactory mucosa, brainstem, cerebellum, and cerebrum[30].

Several researches discuss that entry of SARS-CoV-2 into the central nervous system occurs through its interaction with ACE2[31,32,33]. Vogrig et al.[34] suggested that intracerebral bleeding occurs via attaching of the virus to ACE2 situated on the exterior of endothelial and arterial smooth muscle cells potentially harming cerebral arteries, resulting in the separation or bursting of arterial walls, leading to hemorrhage. Elevated quantities of D-dimer[35-38], fibrinogen, factor VIII[39], von Willebrand factor[40], and tissue factor[41] have been linked to SARS-CoV-2 infection. Szegedi et al.[17] found that elevated D-dimer levels were prevalent, with a median of 3250 ng/mL (IQR: 1140–10,000 ng/mL), and admission fibrinogen levels indicated slight elevation, indicative of systemic inflammation (median: 5.3 g/L, IQR: 4.63–7.39 g/L). Prothrombin time and CRP deviations were common. Remarkably low platelet counts were not documented among reported cases, with counts remaining either within the normal range or only mildly decreased.

According to Bikdeli et al.[42] COVID-19 treatments can interact with oral antiplatelet and anticoagulant agents. Notably, lopinavir/ritonavir, a protease inhibitor used for COVID-19, could impact the effectiveness of commonly administered oral antiplatelet agents such as clopidogrel and ticagrelor due to its inhibition of CYP3A4 metabolism. Prasugrel might be considered as an alternative. Remdesivir, another investigational COVID-19 drug, induces CYP3A4, but dose adjustments for antiplatelet agents are currently not advised. Parenteral antiplatelet agents like cangrelor and glycoprotein IIb/IIIa inhibitors seem unaffected. Concerning anticoagulants, lopinavir/ritonavir could influence dosages of vitamin K antagonists, apixaban, and betrixaban, necessitating potential adjustments, while edoxaban and rivaroxaban should not be combined. Tocilizumab, an IL-6 inhibitor, enhances CYP3A4 expression but doesn’t currently require anticoagulant dose changes. These interactions hold relevance for various clinical indications beyond venous thromboembolism and acute coronary syndrome.

## CONCLUSION

The studies consistently suggest that COVID-19 patients have an increased risk of experiencing cardiovascular and cerebrovascular thromboembolic events. This elevated risk is seen in various conditions such as ischemic heart disease, stroke, and serious cardiac arrhythmias. The long-term burden of thromboembolic events among post-COVID-19 patients is a significant concern. Understanding the persisting impact of these events on patient health, including quality of life, functional limitations, and ongoing medical management, is crucial. The variability in thromboembolic event rates across different medical sites or cohorts raises questions about the impact of healthcare infrastructure, demographics, and other factors. Discussing potential reasons for these variations and their implications for patient care and resource allocation could be insightful. Drawing parallels and differences with other infectious diseases can provide valuable insights into the unique characteristics of COVID-19 and its effects on cardiovascular and cerebrovascular health. The findings of these studies have implications for public health policies and clinical guidelines. Discussing how these findings can inform vaccination strategies, post-COVID-19 care, and surveillance for thromboembolic events can contribute to better healthcare planning. While the studies provide valuable insights, there are likely still many aspects of the relationship between COVID-19 and thromboembolic events that remain unexplored. Discussing potential avenues for future research, including large-scale longitudinal studies and mechanistic investigations, can help guide further understanding of this complex interaction and potential therapeutic targets.

| **Study No.** | **Author Name** | **Sample Size** | **COVID 19 patients** | **Mean Age** | **Male %** | **Cardiovascular events reported** | **Cerebrovascular events reported** |
| --- | --- | --- | --- | --- | --- | --- | --- |
| **1** | **Xie et al.[11]** | 11,650,818 | 153,760 | 63.38 | 90.26 | Atrial fibrillation, Sinus tachycardia and bradycardia, Ventricular arrhythmia, Atrial flutter, Pericarditis, Myocarditis, Acute coronary disease, MI, Ischemic cardiomyopathy, Angina, Heart failure, Non-ischemic cardiomyopathy, Cardiac arrest, Cardiogenic shock Inflammatory heart disease, Ischemic heart disease | Stroke, TIA |
| **2** | **Ortega-Paz et al.[12]** | 4,427 | 3,578 | 60.36 | 53.90 | MI, Cardiovascular death, cardiac arrhythmia, MACE, Adverse cardiovascular event | Ischemic Stroke, Hemorrhagic Stroke, TIA |
| **3** | **Ward et al.[13]** | 763,909 | 417,975 | 52.47 | 36.74 | MI | Stroke |
| **4** | **Siegler et al.[14]** | 14,483 | 14,483 | NA | NA |  | Acute Ischemic Stroke, Intracranial Hemorrhage |
| **5** | **Sabayan et al.[15]** | 18,407 | 15 | 65 | NA |  | Acute Ischemic Stroke, |
|  |  |  |  |  |  |  | Subarachnoid Hemorrhage |
| **6** | **Dhamoon et al.[16]** | 277 | 105 | 66.40 | 52.35 |  | Stroke |

## 1. Contributors

AA made a major contribution to the article, such as the conception of the work and collection of data for the work, correction, tables, and figures editing, and drafted the manuscript from introduction to conclusion. BP contributes to collecting data, double checks for possible errors, and drafting the introduction and method section. PD participates in selecting data, checking for duplicated data, checking for possible errors, and participating in the drafting of method sections and tables. MG participates in checking for data collection, references, and drafting the result section and discussion. MGH participates in drafting discussions, data collection, checking for possible errors, and providing suggestions. RU contributes to abstract drafting, discussion editing, data collection, and checking for possible errors. PD participates in editing the abstract, providing. Suggestions, data collection, figure editing, and title modification. MG and MGH participates in data collection, checks for any possible errors, and drafts conclusions. RU participates in data collection and abstract editing, ensuring all guidelines are met, and drafts limitation sections. AA Participates in generating ideas, providing suggestions, title modification, corrections, revising the manuscript, and drafting the introduction, method, and conclusion. All authors read and approved the final manuscript.

## 2. Funders

No funds, grants, or other support was received.

## 3. Prior presentations

This paper has not been presented anywhere else.

## Data Availability

All data produced in the present study are available upon reasonable request to the authors

